# A Systematic Evaluation of MRI Normalization for Multi-Site Radiomics-Based Disc Degeneration Classification

**DOI:** 10.64898/2026.01.13.26343807

**Authors:** L. Tugan Muftuler, Alexander Drobek, John D. Bukowy, Kathryn Duwe, Vamsi Sudersanam, Joelle Harrington, Eric Van Zant, Savannah R. Duenweg, Dattesh D. Shanbhag

## Abstract

**Objective:** Automated grading of intervertebral disc degeneration from MRI is clinically important but challenging due to inter-rater variability in the Pfirrmann classification and variations in signal intensity and contrast across MRI scanners. Automated approaches may require image normalization to harmonize data from different scanners and ensure robust predictions. Although radiomics-based disease classification offers advantages in explainability and efficiency over deep learning, the effect of image normalization on feature stability and classification performance remains unclear. This study systematically investigated eight MRI normalization pipelines for disc classification and introduced a normalization-robust, interpretable grading tool with performance comparable to human raters.

**Methods:** 270 T2-weighted spine MRIs from different scanners and field strengths were used for training, testing, and validation. Repeat scans from nine subjects enabled assessment of radiomic feature reproducibility. Features were extracted from whole-disc volumes and evaluated across eight normalization pipelines and an unnormalized control. Feature selection prioritized both classification relevance and scan-rescan reproducibility. XGBoost was used for classification.

**Results:** Normalization improved feature reproducibility, but classification performance remained statistically equivalent across all pipelines, indicating robustness to scanner variations. The XGBoost model showed strong agreement with expert grading (Concordance Correlation Coefficient = 0.77), comparable to reported inter-rater agreements. Key features reflected nucleus pulposus desiccation and structural disorganization, supporting biological interpretability. A practical normalization decision framework is presented.

**Conclusion:** Radiomics-based disc degeneration grading is robust to MRI normalization strategy, supporting deployment across different scanners. Its reproducibility, interpretability, and multi-site validation position it as a practical alternative to subjective grading and black-box deep learning.

## INTRODUCTION

Low back pain is a significant health issue with substantial psychosocial and economic consequences ^1^. The degeneration of intervertebral discs is implicated in the majority of chronic low back pain cases, which can lead to various clinically recognizable conditions such as disc herniation, spinal stenosis, and spondylolisthesis ^2-4^. Despite the central role of disc degeneration in spinal pathology, its clinical assessment remains subjective.

Precise and reproducible assessment of disc degeneration has direct implications for patient management ^5^. Patients with mild-to-moderate degeneration often stabilize or respond to conservative therapies, whereas advanced degeneration usually correlates with persistent pain and functional impairment ^6, 7^. Objective, quantitative biomarkers of degeneration could enable prognostic stratification by identifying patients likely to remain stable versus those at risk of progression, and thereby support targeted follow-up intervals and long-term planning. Accurate grading also informs the choice between non-surgical and surgical management, including considerations of adjacent segment disease, implant selection for arthroplasty, and the need for supplemental fixation to avoid fusion complications ^8-13^.

The widely accepted standard for grading disc degeneration is Pfirrmann classification, which grades disc degeneration on a five-point ordinal scale using visual assessment of discs in the mid-sagittal section of T2 weighted MRIs ^8^. But wide variations in appearance of degenerating discs often lead to uncertainty and disagreements between raters, introducing uncertainty into clinical and research decisions alike. There is therefore a clear and unmet need for an objective, reproducible, and clinically deployable grading tool.

Several groups have sought to address this need through automated grading by training Convolutional Neural Networks (CNN) on T2 weighted MRIs ^14, 15^. While CNNs can achieve high accuracy, they are large, data-hungry models that require extensive training sets to learn the full variation in disc shape and signal intensity, and their internal representations remain opaque, which is a significant barrier to clinical trust and regulatory approval. Natalia et al. proposed a more interpretable approach combining automated disc height calculation with color correlogram features across six classifiers ^16^. McSweeney et al. recently applied radiomics features to Pfirrmann grading using data from a single 1.5T scanner ^17^.

Radiomics-based approaches are particularly attractive because they reduce thousands of voxel-level measurements into a compact, mathematically defined feature set that can be linked to tissue microstructure and pathophysiology. However, prior work by McSweeney et al. has several limitations: data were acquired on a single scanner, limiting generalizability; features were extracted only from the mid-sagittal slice, increasing sensitivity to spinal curvature; and feature selection based on correlation with expert grades risks propagating rater bias into the model. Critically, no existing study has systematically evaluated how image normalization for multi-site harmonization affects radiomic feature stability and downstream classification performance in disc degeneration.

Without this understanding, researchers and clinicians cannot make informed decisions about preprocessing pipelines, and the reproducibility of radiomic biomarkers across institutions remains uncertain. Conventional MRI lacks standardized intensity scaling, and contrast characteristics vary substantially across scanner manufacturers, field strengths, and acquisition protocols. These variations can introduce systematic feature bias unrelated to underlying tissue pathology. Although normalization methods aim to harmonize images to a common reference, their comparative effectiveness in musculoskeletal radiomics has not been established.

It is plausible that some normalization approaches improve feature reproducibility without improving, or even degrading, classification performance if biologically relevant signal is suppressed along with scanner-induced variability. Conversely, if radiomic features are inherently robust to scanner variation, complex normalization may be unnecessary. To address these questions, this study presents a controlled, head-to-head benchmark using repeat-scan data to quantify reproducibility.

The primary objective of this study was to conduct the first systematic benchmark of MRI normalization strategies for spinal disc radiomics, using a multi-site dataset and repeated scans to quantify the joint effect of normalization on feature reproducibility and classification performance. Our central hypothesis was that normalization would enhance radiomic feature stability, yet the classification performance of a well-chosen radiomic feature set would remain robust across normalization strategies. This finding would have significant implications for clinical deployment in diverse scanner environments.

To achieve this, (1) we used data from different scanners and field strengths; (2) extracted radiomic features from the whole disc volume, eliminating sensitivity to mid-sagittal slice selection and spinal curvature; (3) selected features based jointly on classification significance and scan-rescan reproducibility, reducing the propagation of rater bias; (4) benchmarked eight normalization pipelines with a formal equivalence analysis of their effect on classification; and (5) postulated about associations between discriminative radiomic features and disc pathophysiology, providing a biologically interpretable basis for automated grading.

These contributions position the present work not only as a grading tool but also as a methodological reference for the rapidly expanding field of musculoskeletal radiomics. They provide evidence-based guidance on selecting normalization strategies and establish the biological plausibility of radiomic biomarkers for disc degeneration.

## METHODS

### MRI data

270 T2-weighted spine MRIs were used for training, testing, and validation. These MRIs were acquired from four different 3T and three 1.5T scanners from three different manufacturers. 208 of these MRIs were obtained from an online repository (https://zenodo.org/records/10159290), while 62 were acquired at our site using a 3T GE Signa Premier MRI system (Waukesha, WI USA). The pulse sequence parameters and spatial resolutions varied across the scanners. Most of the repository data had 3.3mm slice thickness and 0.6mm in-plane resolution and our site data had 1 mm isotropic resolution. The diversity in the dataset ensured that both the training and test sets captured diverse imaging characteristics and variations inherent to different MRI acquisition protocols.

Nine participants from our site were scanned twice on different days, which were used for analyzing reproducibility of features. The five lumbar discs were graded by experienced raters at respective sites using Pfirrmann classification ^8^.

The study conducted at our site was approved by the Institutional Review Board (IRB), and participants provided written consent. The repository data documentation stated that they had IRB approval for their study.

The repository data included labels for discs and vertebral bones, along with a spreadsheet containing radiological evaluations. Binary masks for lumbar discs in the local site data were automatically segmented using nnU-Net. ^18^. To achieve this, we trained a full-resolution 3D model in a 5-fold cross-validation over the training cases. Training was done using the full repository data and 20 scans from our site, which were labeled by the GE Healthcare research team. Each fold was split into 167 training and 42 validation cases. Average pseudo-DICE values for the five lumbar discs across five folds were [0.90, 0.89, 0.86, 0.83 and 0.83]. Further details of nnU-Net models can be found in the paper and their online documentation.

### Radiomic Features

Radiomics refers to computational algorithms that extract quantitative features from medical images ^19,20^. If the features are sensitive and specific to disease-related changes, one can obtain a precise and unbiased assessment of the disease. Since a degenerating disc exhibits both shape deformations and regions of low signal intensity in T2 weighted MRI scans, a limited set of shape and gray level features might be able to quantify the extent of disc degeneration. More importantly, radiomics will not be affected by the variations in location of low intensity regions and shape deformations between discs of the same grade. Thus, it provides a simple and robust feature set for training a classification model. As a result, a much smaller model and training data set should be sufficient compared to the conventional Deep Learning (DL) approaches. Additionally, radiomics features represent physical quantities, which provides explainability to the outcome of the classification model.

Pyradiomics software ^20^ was used to calculate a set of features from each lumbar disc. The software calculates a set of of first-order statistics, shape descriptors, and three texture classes. We initially included the first order statistics, 2D and 3D shape features, Gray Level Co-occurrence Matrix (GLCM), Gray Level Dependence Matrix (GLDM), Gray Level Run Length Matrix (GLRLM), Gray Level Size Zone Matrix (GLSZM), and Neighbouring Gray Tone Difference Matrix (NGTDM) from the texture classes. Then, stable features from repeated scans were determined separately for each normalization method and classification used only those features for prediction. Although Pfirrmann classification requires only viewing the mid-sagittal slice for evaluation, we used the entire volume of the disc for feature calculations which should provide a more accurate representation of the degenerative changes.

### Image data normalization

Voxel values in conventional T2-weighted MR images do not represent a physical quantity and are arbitrarily scaled. It is a common practice to normalize MR images to account for this variability before performing calculations on multi-site data. Therefore, we explored whether normalization was necessary to improve the test-retest consistency of radiomic features by reducing systematic inter- and intra-subject bias, and whether a specific normalization method performed better than others. The *normalization* setting in Pyradiomics software itself was set to *False* to avoid any interference. A control condition without any normalization was used as a baseline and eight normalization methods were tested:

1. Global histogram equalization: Apply histogram equalization to the entire image.
2. Local histogram equalization: Apply local histogram equalization using adaptive histogram equalization ^21^
3. Mask normalization: I(x,y,z) / VB_mean_; Where I(x,y,z) is the entire image and VB_mean_ is the mean value of the voxels in vertebral bodies.
4. z score normalization: (I(x,y,z) - VB_mean_)/ VB_std_; where VB_std_ is the standard deviation in the vertebral bodies.
5. Mask normalization (4) followed by local histogram equalization (3).
6. Mask normalization (4) followed by global histogram equalization (2).
7. z score normalization (5) followed by global histogram equalization (2).
8. z score normalization (5) followed by local histogram equalization (3).

For the histogram equalization methods, the intensity values are scaled between 0-1 for the equalization procedure, then scaled back into their original values. Each subject’s vertebrae were used as reference regions of interest (ROIs) for the mask-based methods.

### Test-retest bias of radiomics features by preprocessing approach and subsequent model training

Nine participants underwent two MRI scans on different days, which were used for this analysis. The stability analysis pipeline assessed potential systematic test–retest bias in radiomics features across repeated scans for each normalization method separately by testing whether the distribution of feature deltas (differences between first and repeated scans) differed from zero using a one-sample t-test. Features with ≥ 0.2 were interpreted as lacking sufficient evidence of mean test–retest shift and were therefore retained as candidate stable features for purposes of subsequent classification modeling. A text file containing the retained features was generated for each normalization method and these were used for respective classification model development.

### Classification

The MRI scans were arranged into identical training and test splits across all normalization conditions, allowing model comparisons to be made on a case-matched basis. Extreme Gradient Boosted decision trees ^22^ (XGBoost) was then used to classify the Pfirrmann score. Gradient boosting is a machine learning technique that builds predictive models in a sequential manner by combining the outputs of multiple weak learners, typically decision trees. XGBoost also provides feature importance metrics based on the learned tree structure. The mean and standard deviation of each importance metric are then calculated.

XGBoost hyperparameters were optimized using a grid search performed on the unnormalized dataset using GridSearchCV tool in scikit-learn. The parameters were then applied to all remaining models. Nine models, one per normalization method, were trained using their respective retained feature sets following test-retest stability screening. Model performance was compared using a repeated cross-validation design to evaluate whether any normalization method yielded a statistically superior result. F1 score was used as the primary metric, which balances precision and recall to provide a robust measure of predictive performance in the presence of class imbalance. It is calculated using F1 = 2*TP/(2*TP+FP+FN), where TP is true positives, FP is false positives and FN is false negatives. Receiver operating characteristic (ROC) curves were also generated for each model, and the area under the curve (AUC) was used as a complementary performance metric. Cross-validation was performed to estimate 95% confidence intervals for the ROC AUC values, enabling statistical comparison across normalization methods. 75% of the data was used for the training. Then, a final evaluation of the model’s performance is done using the unseen 25% of the data. Since the degeneration scores were not evenly distributed across all discs studied here, class based stratified splitting was applied after subject level stratification. This ensures that the class distribution is similar for each split. The distribution of degeneration grades are shown in Fig.1.

**Fig. 1.**
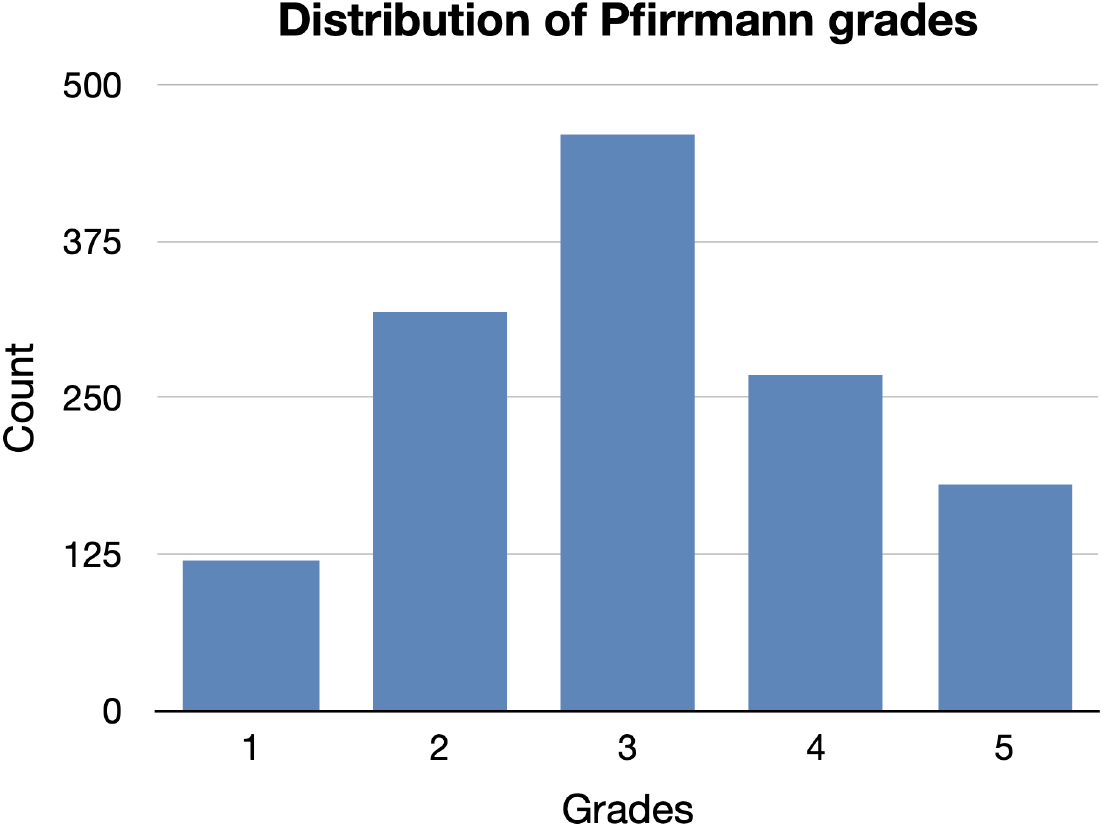
Distribution of Pfirrmann grades in the dataset used for this study.

## RESULTS

### XGBoost predictions

Hyperparameter optimization using GridSearchCV yielded the following values: maximum depth of 12, learning rate of 0.1, number of estimators of 50, 0.8 subsampling, and a regularization weight (alpha) of 0.1.

Fig. 2 presents the mean F1 scores for each preprocessing method. Gray bars represent F1 scores from 5-fold cross-validation, while red bars show F1 scores from an independent test set that was held out and never used during training or validation. The best F1 score was 0.65 for the unseen test data. The Concordance Correlation Coefficient (CCC) between the raters and the prediction was 0.77 for the *Local HE* case, which had one of the highest F1 scores for the test case. Fig. 3 depicts ROC curves that illustrate the classification performance of eight XGBoost models. These models used radiomics features derived from image sets that underwent eight different preprocessing methods. For comparison, a control case is also included, which was not subjected to any preprocessing. Although the mean F1 scores were similar across all methods tested, the *mask normalization with global histogram equalization* and the *z-score* methods had the lowest mean F1. The *mask normalization* and the *control case* performed slightly better than others, and the *mask normalization* had the lowest standard deviation. Interestingly, *control, local histogram equalization* (HE), *global HE* and *z-score* methods performed much better with unseen test data.

**Fig. 2.**
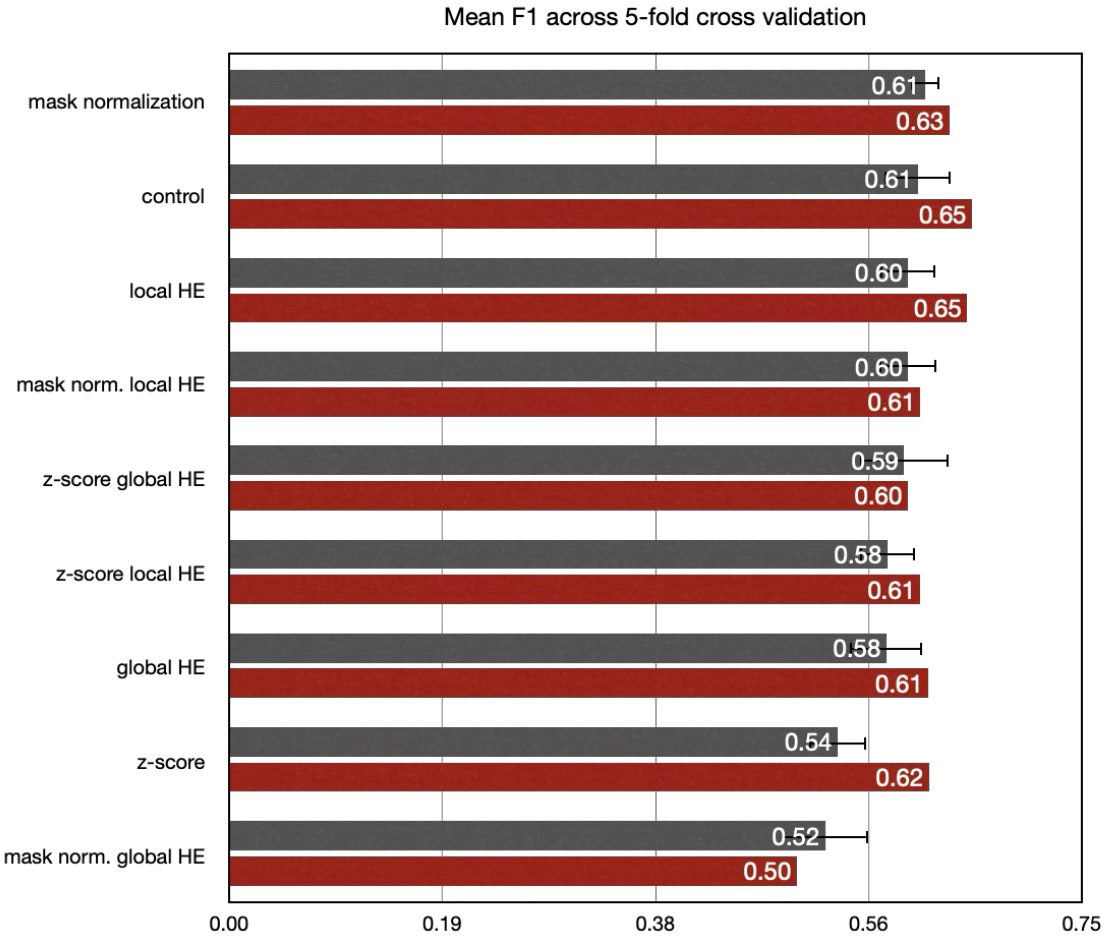
F1 scores for the classification performance from the eight normalization methods and the control case with no normalization. Gray bars show mean and standard deviation of F1 from 5-fold cross validation. Red bars show F1 scores from test data. (HE: histogram equalization).

**Fig. 3.**
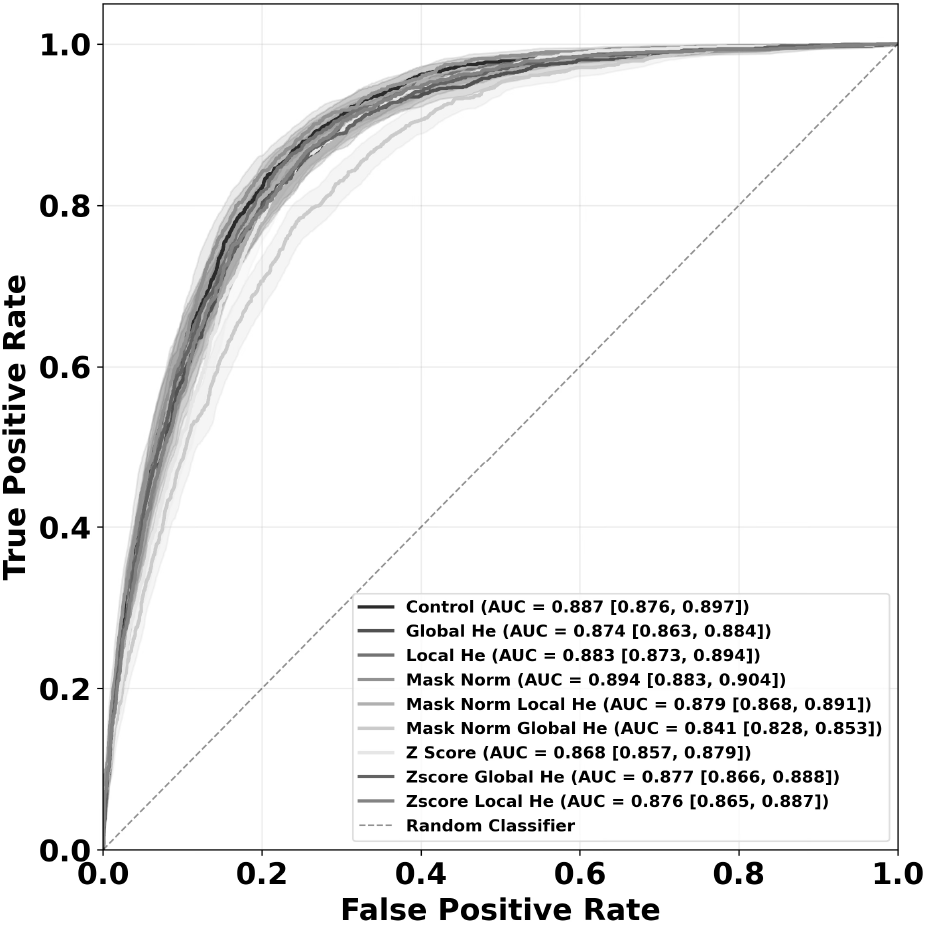
ROC curves with 95% confidence intervals for all preprocessing methods.

These plots illustrate the overall performance of classification for all five degeneration grades. However, when we examined prediction accuracy by grade, the method with the highest accuracy differed across grades. Table 1 presents the AUC values for each preprocessing method for the prediction of each Pfirrmann grade. The accuracy of prediction varied for each grade with different methods. But the values are comparatively close, except for the *mask+Global HE* method.

**Table 1.** AUC for each preprocessing method for predicting each grade. Highest AUC for each grade is underlined and shown in bold fonts. (HE: histogram equalization)

| Preprocessing | AUCs |  |  |  |  |
| --- | --- | --- | --- | --- | --- |
|  | Grade-1 | Grade-2 | Grade-3 | Grade-4 | Grade-5 |
| control | 0.91 | 0.865 | 0.838 | <u>**0.891**</u> | 0.969 |
| Global HE | 0.874 | 0.867 | 0.804 | 0.851 | 0.946 |
| Local HE | 0.916 | 0.849 | <u>**0.856**</u> | 0.889 | 0.961 |
| mask | 0.919 | 0.853 | 0.827 | 0.883 | 0.934 |
| z-score | <u>**0.923**</u> | 0.853 | 0.806 | 0.866 | 0.954 |
| mask+local HE | 0.891 | 0.84 | 0.846 | 0.866 | 0.932 |
| mask+global HE | 0.79 | 0.803 | 0.726 | 0.829 | 0.925 |
| z-score+global HE | 0.823 | <u>**0.875**</u> | 0.816 | 0.847 | 0.961 |
| z-score+local HE | 0.894 | 0.856 | 0.821 | 0.88 | <u>**0.972**</u> |

Fig.4 shows two T2 weighted MRIs from two participants with predicted Pfirrmann grades annotated next to each disc.

**Fig. 4.**
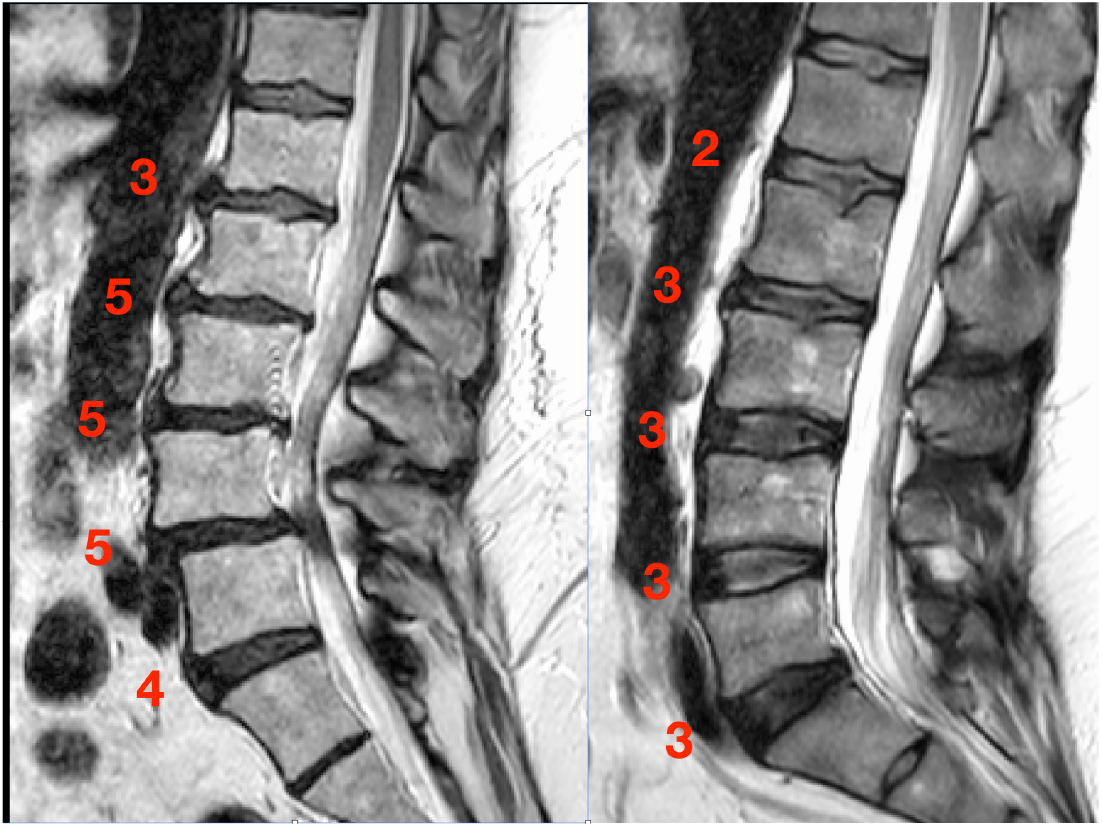
Two T2 weighted lumbar spine images with Pfirrmann scores predicted by XGBoost annotated next to each disc.

### Feature stability

Although image normalization did not improve prediction accuracy noticeably, the number of stable features increased substantially (Fig.5). The number of stable features was 39 without without normalization but increased up to 96 when histogram equalization is introduced.

**Fig. 5.**
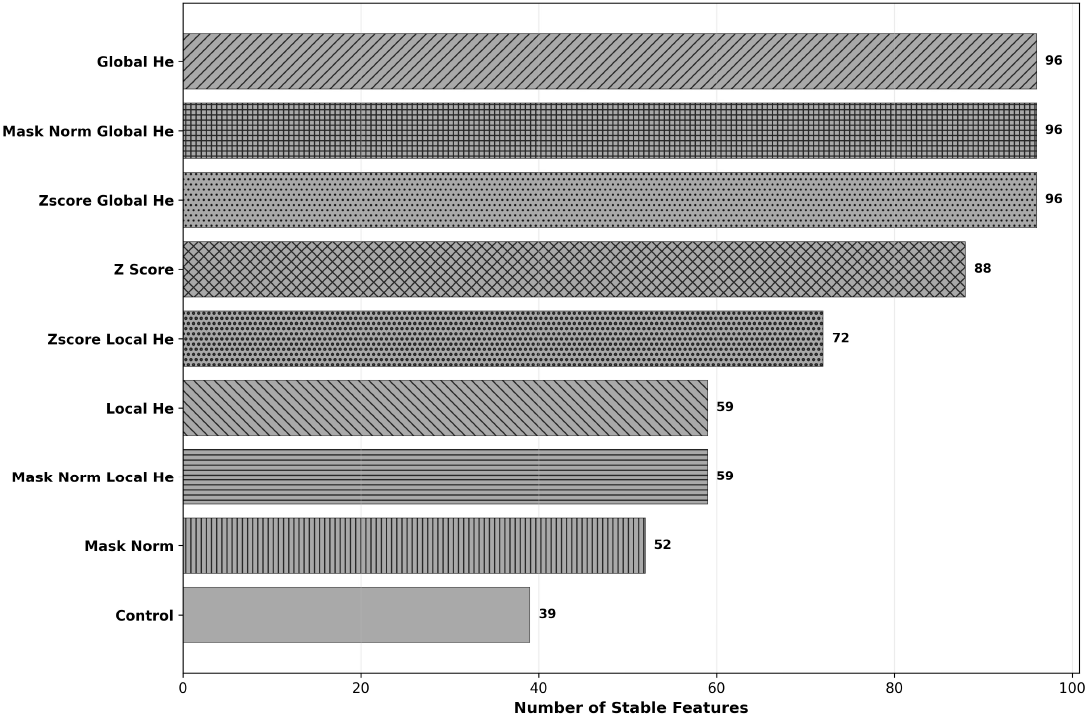
The total number of stable features estimated from repeated MRIs using the normalization methods and the control case.

### Feature importance

XGBoost Feature Importance is calculated using its default gain method, which measures the average improvement in the model’s objective function when a feature is used for splitting. A Pareto front was constructed to visualize the relationship between *feature appearance rate* and model-derived *importance* (Fig.6). For each feature, *feature appearance rate* was defined as the number of normalization methods in which it was retained (i.e., it was stable in repeated scans). It is divided by nine (representing all preprocessing methods). This was plotted against its mean importance value from the trained XGBoost models. Average feature importance is calculated by adding the importances of a feature over nine experiments and dividing by the number of experiments. This representation highlights features that are both consistently stable across preprocessing methods and influential in model prediction. The upper right quadrant contains features that were important in prediction when any one of the preprocessing methods was used. Table 2 lists the top ten robust features that consistently appeared in all nine preprocessing methods. Here the rank is defined as the position in sorted importance list. Rank CV is calculated by taking the standard deviation of a feature’s rank across the nine experiments and dividing it by the average rank. A lower rank CV indicates a better feature, indicating that it consistently appears near the top of the importance list and its position varies little across the nine experiments.

**Fig. 6.**
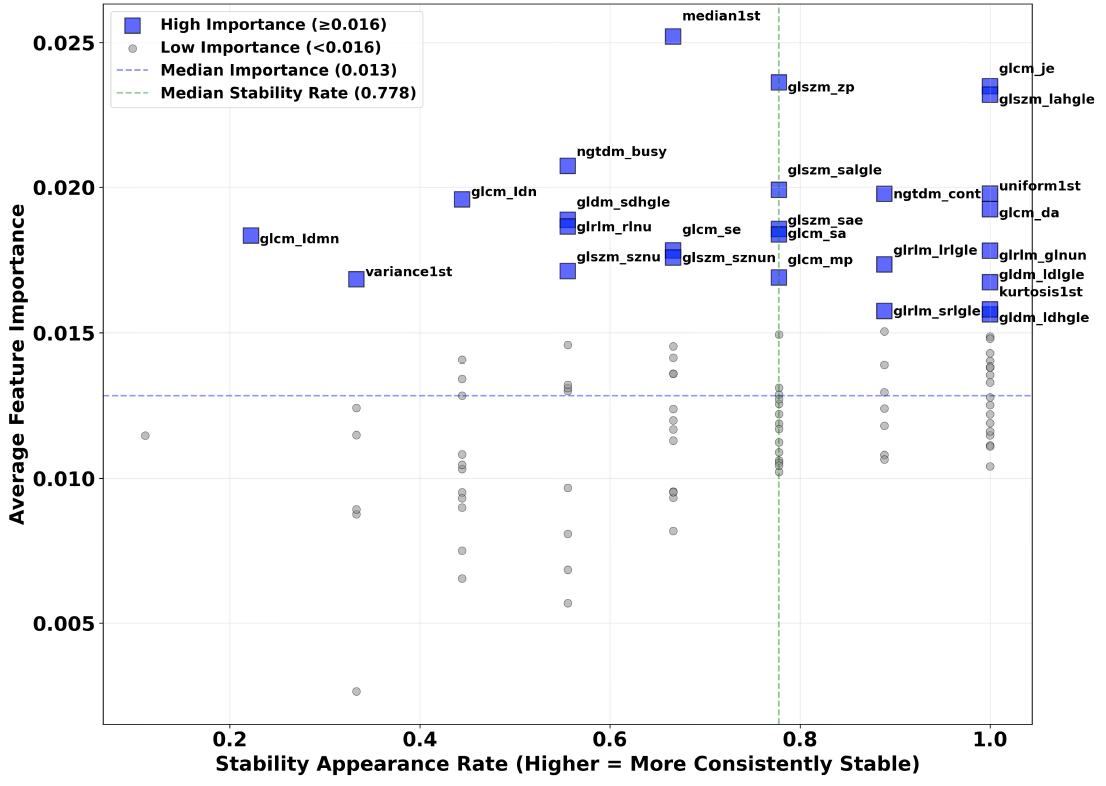
Pareto plot of feature importance and retention following test-retest bias screening in predicting Pfirrmann grades of degenerating discs. The features marked with blue rectangle have high importance in predicting degeneration. The features in the upper right quadrant are both important in prediction and are also more frequently retain post test-retest bias screening across repeated scans. Feature names are abbreviated in this plot. A lookup table for full names are provided in the supplementary data.

**Table 2.** The top ten robust features that consistently appeared in all nine preprocessing methods.

| Radiomics feature | Rank CV |
| --- | --- |
| Kurtosis | 0.35 |
| First order Minimum | 0.42 |
| GLCM JointEnergy | 0.57 |
| GLDM DependenceEntropy | 0.83 |
| GLDM DependenceNonUniformity | 0.39 |
| GLDM DependenceNonUniformityNormalized | 0.42 |
| GLDM LargeDependenceHighGrayLevelEmphasis | 0.89 |
| GLDM LargeDependenceLowGrayLevelEmphasis | 0.55 |
| GLDM SmallDependenceEmphasis | 0.63 |
| GLSZM LargeAreaHighGrayLevelEmphasis | 0.45 |

## DISCUSSION

### Normalization does not improve prediction accuracy

This study addressed three primary objectives: (1) to establish a systematic benchmark of eight MRI normalization strategies for multi-site radiomics-based disc degeneration grading; (2) to demonstrate that radiomics-derived features are resilient to scanner-induced signal intensity and contrast variations; and (3) to identify features that are retained following test-retest bias screening and are important for classification. The central finding of this work is that the proposed pipeline is robust to normalization and enables deployment across different scanners. XGBoost was selected as a well-established, high-performing baseline for structured tabular data, enabling consistent and interpretable comparison across all preprocessing conditions. The inclusion of multi-site MRI data spanning different field strengths and spatial resolutions further strengthens the generalizability of these conclusions.

Predictions derived from unprocessed (control) images and from seven normalization methods achieved comparable agreement with expert raters, with CCC values consistent with those reported in inter-rater reliability studies and prior automated grading approaches ^8, 23-25^. This consistency across normalization conditions, including the no-normalization control, demonstrates that radiomics-based disc degeneration grading is inherently robust to the contrast and signal intensity variability that typically complicates multi-site MRI studies. This resilience is important for clinical deployment, as it implies that elaborate site-specific preprocessing pipelines are not a prerequisite for reliable grading performance. The sole exception was mask normalization combined with global histogram equalization, which consistently degraded prediction quality relative to all other conditions, and should therefore be avoided in radiomics-based spinal imaging workflows.

This finding challenges the general assumption that normalization is a prerequisite for quantitative analysis of multi-site data. While this may be required for other methods, radiomics appear to be more resilient to scanner-to-scanner variations. Our results demonstrate that while normalization does improve feature-level test-retest consistency (by more than two-fold relative to the control condition), this improvement in feature stability does not translate into improved classification performance on the five-grade Pfirrmann scale. This dissociation between feature stability and predictive utility indicates that features that are important for predicting disc degeneration are reproducible without the need for normalization. Investigators applying radiomics in other disease contexts should carefully evaluate whether normalization-induced stability gains yield corresponding gains in downstream task performance.

### Grade-level performance

Prediction accuracy was broadly consistent across Pfirrmann grades for the no-normalization case and the majority of normalization methods. AUC values were marginally higher for Grades 1 and 5 than for intermediate grades, reflecting the well-established challenge of boundary ambiguity between Grades 2, 3, and 4. This ambiguity is not unique to automated methods; it is a known source of inter-rater disagreement in human grading, stemming from the gradual and overlapping nature of degenerative changes across intermediate stages ^26^. Accordingly, some degree of disagreement between XGBoost predictions and expert grades at intermediate stages is expected and does not represent a failure of the proposed pipeline. Rather, it mirrors the inherent subjectivity of the Pfirrmann system itself. Importantly, because the biases introduced by individual expert raters are likely to be distributed approximately randomly across a large, multi-site training dataset, the radiomics-based approach presented here provides a less systematically biased estimate of degeneration stage than any single rater’ s assessment.

### Feature extraction from whole disc volume versus from mid-sagittal

Prior radiomics-based approach to disc grading ^17^, relied exclusively on features extracted from the mid-sagittal slice, thereby mimicking the visual assessment performed by human raters. Our approach extracts features from the entire disc volume, enabling capture of degenerative changes that may be asymmetrically distributed or peripheral to the central sagittal plane, which could be missed by a mid-sagittal approach. This is particularly relevant in patients with mild scoliosis or suboptimal positioning, where the anatomical centre of the disc may not align with the mid-sagittal image plane. Whole-volume segmentation also eliminates the need for per-image identification of the central section of each disc, reducing a potential source of error and operator dependency. Feature reproducibility was assessed using actual repeated MRI acquisitions and paired T-tests to identify features with statistically significant test-retest differences. This is a methodologically more rigorous approach than the image perturbation and Intraclass Correlation Coefficient strategy employed in prior work ^17^, which simulates but does not directly measure true scan-rescan variability

### Biological interpretation of important features

A critical advantage of the radiomics framework over DL approaches such as CNNs or Multi-Layer Perceptrons is the physical interpretability of the features driving predictions. In contrast to the black-box nature of end-to-end DL, each radiomics feature corresponds to a quantifiable property of the image that can be mapped onto known disc biology. The high-importance features identified by XGBoost in this study are not only statistically significant, but they are biologically meaningful in the context of disc degeneration as visualized on T2-weighted MRI.

Specifically, a healthy disc is characterized by high and uniform water content within the nucleus pulposus, which manifests as high signal intensity and high uniformity on T2-weighted images. Features such as *Large Area High Gray Level Emphasis, Uniformity*, and *Energy* therefore take high values in healthy discs and decline progressively as degeneration advances and the nucleus pulposus loses water content and is replaced by fibrocartilage. In early degeneration, this process is spatially localized, producing heterogeneous signal patterns captured by features such as *Gray Level Non-Uniformity* and *Small Area Low Gray Level Emphasis*. As degeneration progresses, these changes extend across the disc, and Kurtosis, which reflects the sharpness of the signal intensity distribution, becomes an increasingly informative discriminator between intermediate and advanced grades. This feature-to-pathophysiology mapping provides an internally consistent biological narrative for the model’s predictions, enhancing clinical trust and facilitating future hypothesis-driven feature engineering

### Clinical application

The clinical utility of this tool lies in its potential to support prognostic stratification and treatment planning. We have previously demonstrated the existence of a threshold beyond which disc degeneration accelerates rapidly ^27^. The probability outputs of the XGBoost classifier, rather than discrete grade predictions alone, could be used to model a patient’s position along this degenerative continuum, providing richer prognostic information than a categorical Pfirrmann grade. For instance, two patients with Pfirrman grade 3 discs may differ substantially in their proximity to the accelerated-degeneration threshold, with direct implications for clinical decision-making. Future work should explore whether baseline radiomics-derived degeneration probability scores predict longitudinal progression, which would elevate this tool from a cross-sectional grading aid to a prognostic biomarker.

### Limitations

The main limitation shared by this and comparable studies is the reliance on Pfirrmann grades, a subjective, ordinal classification system, as the training target. No objective histopathological gold standard for in vivo disc degeneration grading currently exists. Incorporating histopathological information would require the development of a parallel grading framework, which remains an important direction for the field. However, under the assumption that rater-assigned grades approximate a normal distribution around the true degeneration state, the aggregated multi-site, multi-rater dataset used here substantially attenuates systematic individual rater biases. While this study demonstrates its cross-site robustness, it’s crucial to conduct a prospective assessment of its impact on clinical decision-making to determine its clinical value.

## CONCLUSION

This study presents the first systematic benchmark of MRI normalization strategies for radiomics-based intervertebral disc degeneration grading, using a multi-site dataset spanning multiple field strengths and scanner types. The central finding, which shows that radiomics-based predictions are robust to normalization method choice, is not a null result. In fact, it shows that reliable, objective disc grading does not require elaborate site-specific preprocessing, substantially simplifying the path to clinical deployment.

The use of whole-disc-volume features, biologically interpretable high-importance predictors, and reproducibility screening based on true repeated scans collectively represent methodological advances over prior radiomics-based grading approaches. The transparent, feature-driven nature of the pipeline enables direct mapping of model predictions onto known disc pathophysiology, supporting clinical trust and explainability in a way that black-box DL approaches cannot.

Finally, the probabilistic outputs of the classifier offer a foundation for future prognostic stratification research, particularly in relation to the previously identified degenerative threshold beyond which disc deterioration accelerates. Longitudinal validation of whether baseline radiomics scores predict degeneration progression represents the critical next step toward establishing this tool as a true prognostic biomarker.

## Supporting information

detailed list of features used in analysis

## Data Availability

The majority of the data was obtained from online repository (https://zenodo.org/records/10159290). The data acquired at our site are available upon reasonable request to the authors.

## Acknowledgements

We would like to thank research coordinators Amy Nader, Kearnin Van Bortel and Samatha Omidfar for their assistance in this study. The software engineering team from GE healthcare provided additional data for segmentation of discs and vertebral bones to train the nnU-Net.

## Grant Support

This study is supported in part by funds from Advancing a Healthier Wisconsin endowment.

## Conflicts of Interest

The authors declare that there are no relevant conflicts of interest.

## Notes

### Competing Interest Statement

The authors have declared no competing interest.

### Author Declarations

The study conducted at our site was approved by the Institutional Review Board (IRB) of the Medical College of Wisconsin, USA

### Summary of Updates

The main novelty in our work was to explore eight different normalization methods to harmonize data from multiple sites and scanners. The previous version did not emphasize that and focused on prediction of disc degeneration. The revised version primarily focuses on how radiomics feature stability and prediction accuracy is affected by different normalization methods. The title, abstract, introduction and discussion is revised to highlight this novel component of our work.

