## Supplementary material for "A Systematic Evaluation of MRI Normalization for Multi-Site Radiomics-Based Disc Degeneration Classification": detailed list of features used in analysis

Abbreviated feature names

| Feature name | Abbreviation |
|---|---|
| firstorder_10Percentile | 10percentile |
| firstorder_90Percentile | 90percentile |
| firstorder_Energy | energy1st |
| firstorder_Entropy | entropy1st |
| firstorder_InterquartileRange | iqr1st |
| firstorder_Kurtosis | kurtosis1st |
| firstorder_Maximum | max1st |
| firstorder_Mean | mean1st |
| firstorder_MeanAbsoluteDeviation | mad1st |
| firstorder_Median | median1st |
| firstorder_Minimum | min1st |
| firstorder_Range | range1st |
| firstorder_RobustMeanAbsoluteDeviation | rmad_1st |
| firstorder_RootMeanSquared | rms1st |
| firstorder_Skewness | skew1st |
| firstorder_TotalEnergy | total_en_1st |
| firstorder_Uniformity | uniform1st |
| firstorder_Variance | variance1st |
| glcm_Autocorrelation | glcm_ac |
| glcm_ClusterProminence | glcm_cp |
| glcm_ClusterShade | glcm_cs |
| glcm_ClusterTendency | glcm_ct |
| glcm_Contrast | glcm_cont |
| glcm_Correlation | glcm_corr |
| glcm_DifferenceAverage | glcm_da |
| glcm_DifferenceEntropy | glcm_de |
| glcm_DifferenceVariance | glcm_dv |
| glcm_Id | glcm_Id |
| glcm_Idm | glcm_Idm |

| Feature name | Abbreviation |
|---|---|
| glcm_Idmn | glcm_Idmn |
| glcm_Idn | glcm_Idn |
| glcm_Imc1 | glcm_Imc1 |
| glcm_Imc2 | glcm_Imc2 |
| glcm_InverseVariance | glcm_iv |
| glcm_JointAverage | glcm_ja |
| glcm_JointEnergy | glcm_je |
| glcm_JointEntropy | glcm_entropy |
| glcm_MaximumProbability | glcm_mp |
| glcm_MCC | glcm_mcc |
| glcm_SumAverage | glcm_sa |
| glcm_SumEntropy | glcm_se |
| glcm_SumSquares | glcm_ss |
| gldm_DependenceEntropy | gldm_de |
| gldm_DependenceNonUniformity | gldm_dnu |
| gldm_DependenceNonUniformityNormalized | gldm_dnun |
| gldm_DependenceVariance | gldm_dv |
| gldm_GrayLevelNonUniformity | gldm_glnu |
| gldm_GrayLevelVariance | gldm_glv |
| gldm_HighGrayLevelEmphasis | gldm_hgle |
| gldm_LargeDependenceEmphasis | gldm_lde |
| gldm_LargeDependenceHighGrayLevelEmphasis | gldm_ldhgle |
| gldm_LargeDependenceLowGrayLevelEmphasis | gldm_ldlgle |
| gldm_LowGrayLevelEmphasis | gldm_lgle |
| gldm_SmallDependenceEmphasis | gldm_sde |
| gldm_SmallDependenceHighGrayLevelEmphasis | gldm_sdhgle |
| gldm_SmallDependenceLowGrayLevelEmphasis | gldm_sdlgle |
| glrlm_GrayLevelNonUniformity | glrlm_glnu |
| glrlm_GrayLevelNonUniformityNormalized | glrlm_glnun |
| glrlm_GrayLevelVariance | glrlm_glv |
| glrlm_HighGrayLevelRunEmphasis | glrlm_hglre |

| Feature name | Abbreviation |
|---|---|
| glrlm_LongRunEmphasis | glrlm_lre |
| glrlm_LongRunHighGrayLevelEmphasis | glrlm_lrhgle |
| glrlm_LongRunLowGrayLevelEmphasis | glrlm_lrlgle |
| glrlm_LowGrayLevelRunEmphasis | glrlm_lglre |
| glrlm_RunEntropy | glrlm_re |
| glrlm_RunLengthNonUniformity | glrlm_rlnu |
| glrlm_RunLengthNonUniformityNormalized | glrlm_rlnun |
| glrlm_RunPercentage | glrlm_rp |
| glrlm_RunVariance | glrlm_rv |
| glrlm_ShortRunEmphasis | glrlm_sre |
| glrlm_ShortRunHighGrayLevelEmphasis | glrlm_srhgle |
| glrlm_ShortRunLowGrayLevelEmphasis | glrlm_srlgle |
| glszm_GrayLevelNonUniformity | glszm_glnu |
| glszm_GrayLevelNonUniformityNormalized | glszm_glnun |
| glszm_GrayLevelVariance | glszm_glv |
| glszm_HighGrayLevelZoneEmphasis | glszm_hglze |
| glszm_LargeAreaEmphasis | glszm_lae |
| glszm_LargeAreaHighGrayLevelEmphasis | glszm_lahgle |
| glszm_LargeAreaLowGrayLevelEmphasis | glszm_lalgle |
| glszm_LowGrayLevelZoneEmphasis | glszm_lglze |
| glszm_SizeZoneNonUniformity | glszm_sznu |
| glszm_SizeZoneNonUniformityNormalized | glszm_sznun |
| glszm_SmallAreaEmphasis | glszm_sae |
| glszm_SmallAreaHighGrayLevelEmphasis | glszm_sahgle |
| glszm_SmallAreaLowGrayLevelEmphasis | glszm_salgle |
| glszm_ZoneEntropy | glszm_ze |
| glszm_ZonePercentage | glszm_zp |
| glszm_ZoneVariance | glszm_zv |
| ngtdm_Busyness | ngtdm_busy |
| ngtdm_Coarseness | ngtdm_coarse |
| ngtdm_Complexity | ngtdm_complex |

| Feature name | Abbreviation |
| --- | --- |
| ngtdm_Contrast | ngtdm_cont |
| ngtdm_Strength | ngtdm_strength |
| shape_Elongation | elongation |
| shape_Flatness | flatness |
| shape_LeastAxisLength | least_axis |
| shape_MajorAxisLength | maj_axis |
| shape_Maximum2DDiameterSlice | max2diam |
| shape_Maximum3DDiameter | max3Ddiam |
| shape_MinorAxisLength | min_axis |
| shape_SurfaceVolumeRatio | surface/vol |
